# Second wave of the Covid-19 pandemic in Delhi, India: high seroprevalence not a deterrent?

**DOI:** 10.1101/2021.09.09.21263331

**Authors:** Nandini Sharma, Pragya Sharma, Saurav Basu, Ritika Bakshi, Ekta Gupta, Reshu Agarwal, Kumar Dushyant, Nutan Mundeja, Zeasley Marak, Sanjay Singh, Gautam Kumar Singh, Ruchir Rustagi, S K Sarin

## Abstract

**Background:** We report the findings of a large follow-up community-based serosurvey and correlating it with the COVID-19 test-positivity rate and the case load observed during the peak of the second wave of the Covid-19 pandemic in Delhi, India.

**Methods:** Individuals of age ≥5 years were recruited from 274 wards of the state (population ∼ 19.6 million) during January 11 to January 22’ 2021. A total of 100 participants each were included from all the wards for a net sample size of ∼28,000. A multi-stage sampling technique was applied for selection of participants for the household serosurvey. Anti SARS CoV-2 IgG antibodies were detected by using the VITROS assay (90% Sn, 100% Sp).

**Results:** Antibody positivity was observed in 14,298 (50.76%) of the 28,169 samples. The age, sex and district population weighted seroprevalence of the IgG SARS-CoV-2 was 50.52% (95% C.I. 49.94-51.10) and after adjustment for assay characteristics was 56.13% (95% C.I. 55.49-56.77). On adjusted analysis, participants aged ≥50 years, of female gender, housewives, having ever lived in containment zones, urban slum dwellers, and diabetes or hypertensive patients had significantly higher odds of SARS-CoV-2 antibody positivity.

The peak infection rate and the test positivity rate since October 2020 were initially observed in mid-November 2020 with a subsequent steep declining trend, followed by a period of persistently low case burden lasting until the first week of March 2021. This was followed by a steady increase followed by an exponential surge in infections from April 2021 onwards culminating in the second wave of the pandemic.

**Conclusions:** The presence of infection induced immunity from SARS-CoV-2 even in more than one in two people can be ineffective in protecting the population.

## INTRODUCTION

Monitoring the trends of SARS-CoV-2 through serial serosurveys by identifying the true burden and spread of infection constitute an essential element of the public health response for combating the COVID-19 pandemic [1]. In Delhi, the Indian capital city, three previous rounds of population serosurveys observed the presence of IgG antibodies to SARS-CoV-2 in 28.39% (August 2020), 24.08% (September 2020), and 24.71% (October 2020) [2].

We report the findings of a large follow-up community-based serosurvey and correlating it with the COVID-19 test-positivity rate and the case load observed during the peak of the second wave of the Covid-19 pandemic in Delhi, India.

## METHODS

Study design, participants, and settings: This was a cross-sectional seroepidemiological study among individuals of age ≥5 years recruited from 274 wards of the state of Delhi (population ∼ 19.6 million) during January 11 to January 22’ 2021.

A total of 100 participants each were enrolled from all the wards except the Delhi cantonment and the New Delhi wards due to their disproportionately larger size. This sample size of ∼28,000 was estimated at 99% confidence levels, 1% absolute precision, expected prevalence 25% from the prior serosurvey round [2], design effect of 2, and considering 15% non-response. It was also adequately powered to elicit seroprevalence comparisons at the district level.

The housing settlement types in the state of Delhi are classified as planned colony/urban slum/resettlement colony/unauthorized colony/rural village [3]. Within each ward, the proportion of participants selected from each settlement type was stratified as per their tentatively estimated population size. A multi-stage sampling technique was applied for selection of participants for the household serosurvey using the following steps: (i). Simple Random Sampling to select the sampling areas within each settlement type (ii). Systematic Random Sampling to select the households within the selected sampling areas. (iii). Selection of an individual participant from every selected household using the age-order procedure.

Laboratory procedure: Around 3-4 ml venous blood was collected through venepuncture by a trained phlebotomist or lab technician under all aseptic precautions. The samples were transported and processed at the Clinical Virology Lab, Institute of Liver & Biliary Sciences, New Delhi. Anti SARS CoV-2 IgG antibodies were detected by using the VITROS assay on VITROS 3600 instrument based on Chemiluminescent technology as per the kit literature [4].

Statistical Analysis: the sociodemographic data of the participants collected through a brief interview schedule was entered in Microsoft Excel 2013 and merged with their antibody test results. The data were analysed with IBM SPSS Version 25 (Armonk, NY: IBM Corp) and Stata 14 (StataCorp, USA). The seroprevalence estimates were weighted to match the state demographics by age and sex and reported as proportions with 95% confidence intervals. We adjusted the weighted seroprevalence for the assay characteristics using the Rogan-Gladen estimator, where True (Adjusted) prevalence = Weighted Prevalence + (Specificity – 1) / [Specificity + Sensitivity - 1] [5].

The data estimates of COVID-19 cumulative case burden, recovery and the test positivity rates were obtained from the official state government sources. A p-value < 0.05 was considered statistically significant.

### Ethical considerations

Written and informed consent for adults and assent and parental consent for minor participants was obtained. The study was approved by the Institutional Ethics Committee, Maulana Azad Medical College & Associated Hospitals, 2 BSZ Marg, New Delhi - 110002 F.1/IEC/MAMC/(78/06/2020/No.176).

## RESULTS

A total of 28,169 laboratory samples were successfully processed excluding the 733 samples that were damaged during transit, haemolysis, label mismatch or inadequate blood collection. The non-response rate at the household level was estimated at 18%. Antibody positivity was observed in 14,298 (50.76%) participants. The age, sex and district population weighted seroprevalence of the IgG SARS-CoV-2 antibody was 50.52% (95% CI 49.94-51.10). After adjustment for assay characteristics, the seroprevalence was estimated to be 56.13% (95% CI 55.49-56.77).

The weighted and adjusted seroprevalence (95% C.I) in the districts ranged from the lowest in North District 49.09 (46.69-51.51) to the highest in the South-East District 62.18 (60.12-64.21) (Figure 1).

**Figure 1.**
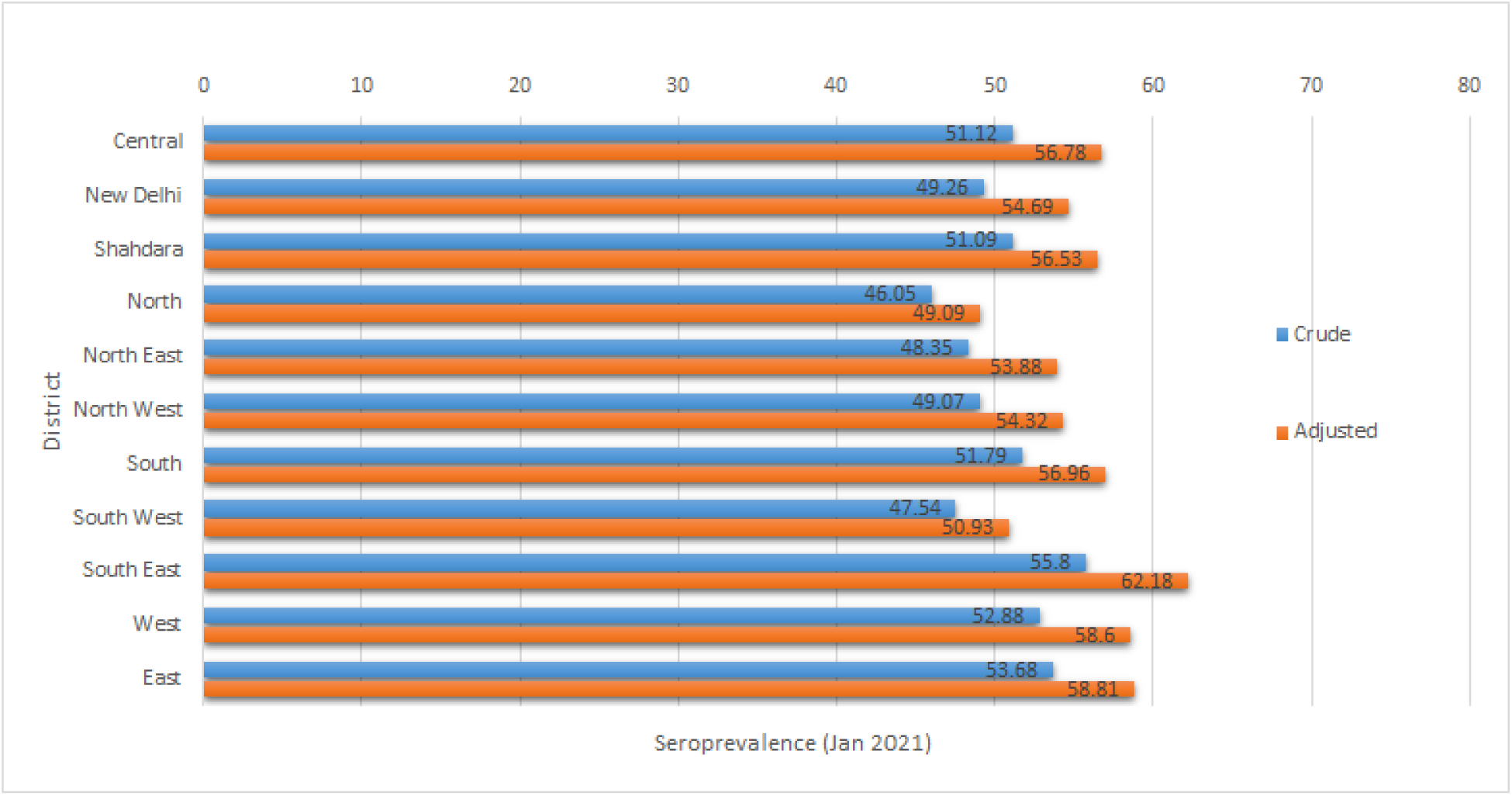
Seroprevalence of SARS-CoV-2 in 11 districts of Delhi, India

The age and gender stratified seroprevalence estimates are reported in Table 1. On adjusted analysis, participants aged ≥50 years, of female gender, housewives, those who had ever lived in containment zones, urban slum dwellers, and diabetes or hypertensive patients, had significantly higher odds of SARS-CoV-2 antibody positivity (Table 2). Furthermore, only 72.3% of the participants with a self-reported past history of COVID-19 diagnosed through either molecular or antigen methods (n=1121) were having detectable IgG antibodies to SARS-CoV-2.

**Table 1.** Seroprevalence stratified by district, age and gender^*^.

| Age | <18 |  | 18 to 49 |  | >50 |  |
| --- | --- | --- | --- | --- | --- | --- |
| District | Male | Female | Male | Female | Male | Female |
| Central | 54.36 | 58.60 | 47.64 | 49.72 | 52.26 | 57.75 |
| New Delhi | 54.86 | 60.13 | 42.26 | 49.72 | 46.55 | 58.39 |
| Shahdara | 52.81 | 54.31 | 47.03 | 49.74 | 50.79 | 60.48 |
| North | 39.47 | 44.51 | 39.53 | 48.99 | 50.88 | 55.21 |
| North East | 50.52 | 47.06 | 47.26 | 50.47 | 44.50 | 46.98 |
| North West | 51.93 | 55.77 | 44.87 | 50.20 | 47.81 | 49.89 |
| South | 49.49 | 53.81 | 50.54 | 54.27 | 45.45 | 54.15 |
| South West | 38.55 | 50.94 | 46.95 | 46.13 | 49.01 | 51.40 |
| South East | 60.58 | 60.07 | 48.58 | 55.44 | 56.71 | 66.08 |
| West | 58.85 | 58.08 | 47.36 | 53.03 | 52.21 | 60.64 |
| East | 53.60 | 49.26 | 52.43 | 51.54 | 54.87 | 64.10 |
| Total | 51.28<br>(49.10,53.45<br>) | 54.30<br>(52.20,<br>56.39) | 46.91<br>(45.76,<br>48.06) | 50.97<br>(50.03,<br>51.91) | 50.35<br>(48.40, 52.29) | 56.72<br>(54.92, 58.51) |

\*All calculations performed on unweighted data

**Table 2.** Distribution of socio-demographic and clinical characteristics with IgG SARS-CoV-2 (N=28,152)^*^

| Variable | N=28152 n, (%) | IgG Seropositive | Unadjusted odds | Adjusted odds |
| --- | --- | --- | --- | --- |
| Age (in Years) |  |  |  |  |
| <18 | 4337 (15.41) | 2274 (52.43) | 1 | 1 |
| 18-49 | 18259 (64.88) | 8996 (49.27) | 0.88 (0.82, 0.94) | 0.89 (0.81, 0.97) |
| ≥50 | 5548 (19.71) | 2983 (53.77) | 1.06 (0.97, 1.14) | 1.03 (0.93, 1.14) |
|  |  |  | <0.001 | <0.001 |
| Gender |  |  |  |  |
| Male | 11861 (42.13) | 5718 (48.21) | 1 | 1 |
| Female | 16291 (57.87) | 8539 (52.42) | 1.18 (1.13, 1.24) | 1.16 (1.10, 1.23) |
|  |  |  | <0.001 | <0.001 |
| Education |  |  |  |  |
| Illiterate | 4958 (17.67) | 2579 (52.03) | 1 | 1 |
| Literate | 9774 (34.84) | 4954 (50.69) | 0.95 (0.89, 1.01) | 0.97 (0.91, 1.05) |
| Up to 10<sup>th</sup> | 7841 (27.95) | 3928 (50.10) | 0.93 (0.86, 0.99) | 0.96 (0.89, 1.04) |
| Graduate and above | 5481 (19.54) | 2744 (50.06) | 0.92 (0.86, 1.00) | 0.95 (0.87, 1.04) |
|  |  |  | 0.140 | 0.6915 |
| Occupation |  |  |  |  |
| Self-employed | 3578 (12.75) | 1799 (50.28) | 1 | 1 |
| Salaried | 6192 (22.07) | 3002 (48.48) | 0.93 (0.86, 1.01) | 0.94 (0.87, 1.03) |
| Daily wager | 2045 (7.29) | 934 (45.65) | 0.83 (0.74, 0.93) | 0.86 (0.77, 0.96) |
| HCW | 1449 (5.17) | 750 (51.42) | 1.06 (0.94, 1.20) | 1.01 (0.89, 1.15) |
| Housewife | 9325 (33.24) | 4888 (52.42) | 1.09 (1.01, 1.18) | 1.02 (0.93, 1.11) |
| Student | 5464 (19.48) | 2833 (51.85) | 1.06 (0.98, 1.16) | 1.03 (0.93, 1.14) |
|  |  |  | <0.001 | 0.0136 |
| Overcrowding |  |  |  |  |
| Present | 9435 (33.65) | 4799 (50.86) | 1.01 (0.96, 1.06) | 1.02 (0.97, 1.08) |
| Absent | 18604 (66.35) | 9402 (50.54) | 1 | 1 |
|  |  |  | 0.606 | 0.401 |
| Total family income |  |  |  |  |
| <20,000 | 15798 (56.47) | 8006 (50.68) | 1 | 1 |
| 20,000-50,000 | 7455 (26.65) | 3829 (51.36) | 1.03 (0.97, 1.09) | 1.03 (0.97, 1.09) |
| >50,000 | 4721 (16.88) | 2331 (49.38) | 0.95 (0.89, 1.01) | 0.94 (0.87, 1.01) |
|  |  |  | 0.101 | 0.0474 |
| Toilet facility |  |  |  |  |
| Individual | 7822 (27.78) | 3934 (50.29) | 1 | 1 |
| Shared | 19269 (68.45) | 9816 (50.94) | 1.03 (0.97, 1.08) | 1.07 (1.01, 1.13) |
| Community | 1060 (3.77) | 507 (47.79) | 0.90 (0.80, 1.03) | 0.92 (0.80, 1.05) |
|  |  |  | 0.103 | 0.0088 |
| Settlement type |  |  |  |  |
| Planned | 9655 (34.47) | 5010 (51.89) | 1 | 1 |
| Resettlement | 1701 (6.07) | 851 (50.03) | 0.93 (0.84, 1.03) | 0.93 (0.84, 1.04) |
| Urban Slum | 8844 (31.58) | 4639 (52.45) | 1.02 (0.97, 1.08) | 1.05 (0.98, 1.11) |
| Unauthorized | 3363 (12.00) | 1662 (49.43) | 0.91 (0.84, 0.98) | 0.91 (0.84, 0.99) |
| Village | 4445 (15.87) | 2019 (45.42) | 0.77 (0.72, 0.83) | 0.78 (0.73, 0.84) |
|  |  |  | <0.001 | <0.001 |
| Containment zone ever lived |  |  |  |  |
| Yes | 1383 (4.93) | 797 (57.63) | 1.35 (1.21, 1.50) | 1.13 (1.00, 1.26) |
| No | 26669 (95.07) | 13408 (50.28) | 1 | 1 |
|  |  |  | <0.001 | 0.045 |
| COVID-19<br>positive ever |  |  |  |  |
| Yes | 1121 (4.01) | 817 (72.88) | 2.72 (2.38, 3.11) | 2.95 (2.55, 3.41) |
| No | 26850 (95.99) | 13345 (49.70) | 1 | 1 |
|  |  |  | <0.001 | <0.001 |
| ILI symptoms |  |  |  |  |
| Yes | 599 (2.13) | 315 (52.59) | 1.08 (0.92, 1.27) | 0.80 (0.67, 0.95) |
| No | 27553 (97.87) | 13942 (50.60) | 1 | 1 |
|  |  |  | 0.336 | 0.010 |
| HTN |  |  |  |  |
| Yes | 805 (2.86) | 444 (55.16) | 1.21 (1.05, 1.39) | 0.91(0.77, 1.06) |
| No | 27347 (97.14) | 13813 (50.51) | 1 | 1 |
|  |  |  | 0.009 | 0.229 |
| DM |  |  |  |  |
| Yes | 785 (2.79) | 472 (60.13) | 1.49 (1.29, 1.72) | 1.32 (1.12, 1.55) |
| No | 27367 (97.21) | 13785 (50.37) | 1 | 1 |
|  |  |  | <0.001 | 0.001 |

\*17 samples had missing questionnaire data; all analysis performed on unweighted data

The infection fatality ratio per 1,000 cases ranged from 0.98 (0.96, 1.00) to 0.98 (0.97, 1.01). The infection to case ratio per 1,000 cases ranged from 56.98 (56.84, 57.12) to 57.31 (57.17, 57.45).

The peak infection rate and the test positivity rate since October 2020 were observed in mid-November 2020 with a subsequent steep declining trend, followed by a period of persistently low case burden lasting until the first week of March 2021. This was followed by a steady increase and subsequently followed by an exponential surge in infections from April 2021 onwards indicating the second wave of the pandemic (Figure 2).

**Figure 2.**
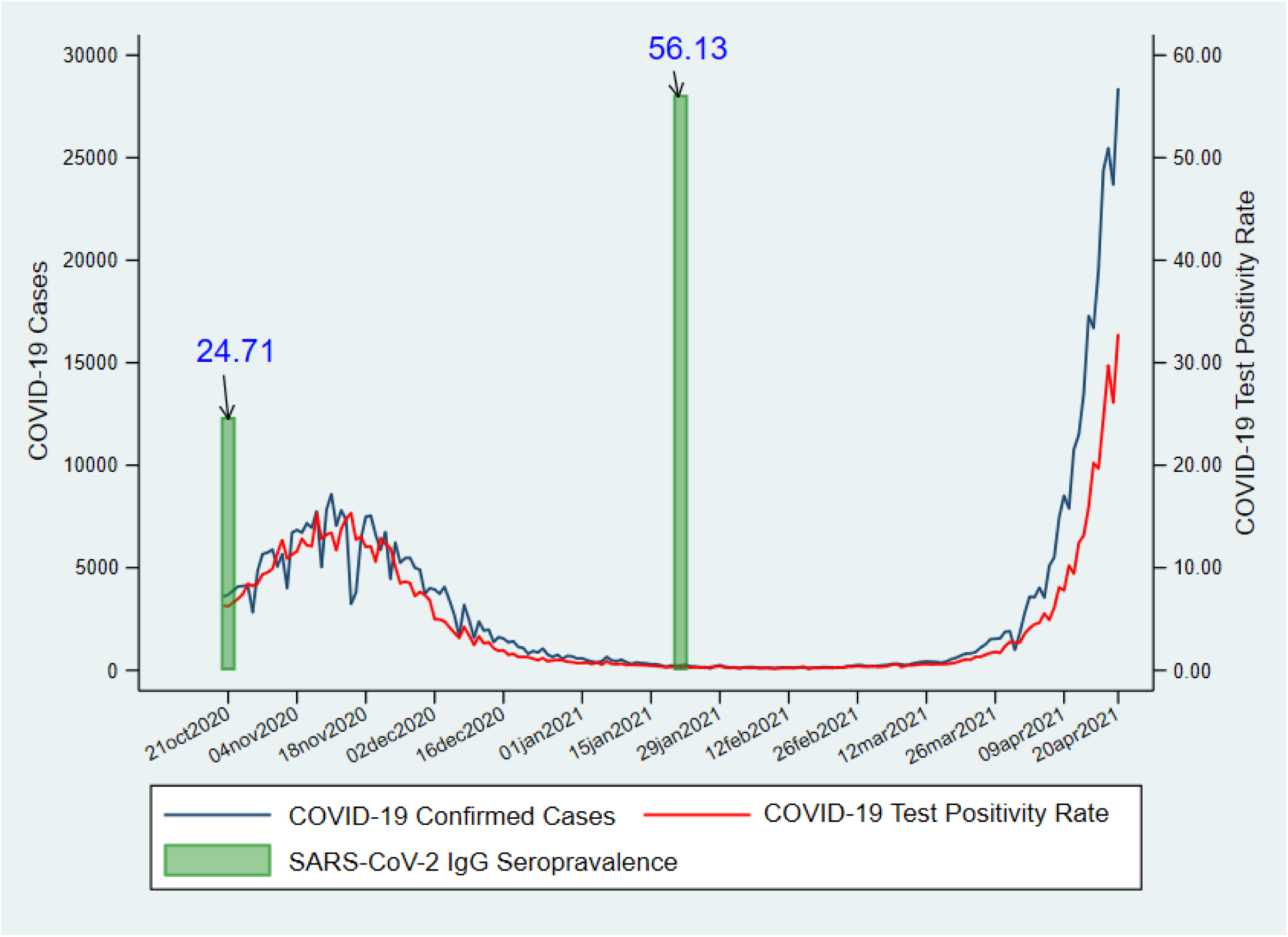
Epidemic curve and seroprevalence of SARS-CoV-2 in Delhi (Oct’2020 – April’2021)

## DISCUSSION

In this serosurvey, a majority of the participants had detectable antibodies to SARS-CoV-2 from past infection with nearly uniform district level trends. In comparison to the three months preceding the survey [2], the antibody seroprevalence in January showed a more than two-times increase, coinciding with a rapid decline in the test positivity rate and the daily new incident cases suggestive of a high population-level immunity. However, the high seroprevalence through natural infection was insufficient to achieve herd immunity and avert the second wave of the pandemic in Delhi when nearly 0.737 million cases including 11,075 deaths were observed in the period from April to May’ 2021 [6]. Previously, the possibility of resurgence of cases despite high seroprevalence had been reported from Manaus in Brazil [7]. Furthermore, current research shows that Covid-19 variants of concern B.1.617.2 (Delta) and B.1.1.7 (Alpha) which have more than two times higher transmissibility and have evolved immune escape mechanisms to potentially bypass antibody response induced from natural infection or vaccine were chiefly responsible for the surge of cases across India including Delhi [8].

In this study nearly one in four participants with a past history of COVID-19 disease were seronegative, a finding consistent with previous rounds where antibodies to COVID-19 were lacking in a substantial proportion of the participants [2]. These findings could be attributed to the growing evidence suggestive of the waning of IgG antibodies especially during asymptomatic and mild illness, although the risk of Covid-19 reinfection in previously infected patients remains low [9, 10].

The present study has some important implications for public health management of the COVID-19 pandemic particularly in densely populated lower middle income countries. First, the presence of infection induced immunity even in more than one in two people can be ineffective in protecting the population from large scale Covid-19 related morbidity and mortality. Second, nearly one in two participants in the <18 age-group were seropositive, which may have implications towards the opening of educational institutions, and reflect no additional risk in children from exposure to the infection. Third, serosurveys at the district level (sub-national and sub-regional level) may be more appropriate in educating public health policy, compared to national and state level estimates, considering the significant inter-district variation in the observed seroprevalence. Fourth, tracking the emergence of potential variants of concern through robust genomic surveillance and associated contact tracing is necessary [11]. Finally, rapid Covid-19 vaccination with the highest possible coverage remains the most feasible means of combating and ending the Covid-19 pandemic. Nevertheless, emerging evidence suggests natural infections confer a significantly more durable and protective immune response against the Delta strain compared to vaccination [9] signifying the need for serial assessment of antibody response to identify waning response and the possible requirement of a booster dose.

In conclusion, a huge surge of Covid-19 cases culminating in a massive second wave occurred in the city of Delhi despite a majority of the population having evidence of past SARS-CoV-2 infection.

## Data Availability

The anonymized dataset would be made available on reasonable request to the corresponding author.

## Data availability

The anonymized dataset would be made available on reasonable request to the corresponding author.

## Acknowledgments

We thank all the district nodal officers for facilitating the data and sample collection.

Ms. Arti Kakkar for her assistance with the data management for this investigation.

